# Clinical application of monitoring indicators of female dancer health, including application of artificial intelligence in female hormone networks

**DOI:** 10.1101/2021.09.27.21264119

**Authors:** Nicola Keay, Martin Lanfear, Gavin Francis

**Affiliations:** Department of Sport and Exercise Sciences, Durham University, United Kingdom; Head of Performance Medicine, Scottish Ballet; Science4Performance, London, United Kingdom

## Abstract

**Objectives:** The purpose of this study was to assess the effectiveness of monitoring professional female dancer health with a variety of subjective and objective monitoring methods, including application of artificial intelligence (AI) techniques to modelling menstrual cycle hormones and delivering swift personalised clinical advice.

**Methods:** Female dancers from a ballet company completed a published online dance-specific health questionnaire. Over the study period, dancers recorded wellbeing and training metrics, with menstrual cycle tracking and blood tests. For menstrual cycle hormones AI-based techniques modelled hormone variation over a cycle, based on capillary blood samples taken at two time points. At regular, virtual, clinical interviews with each dancer, findings were discussed, and personalised advice given.

**Results:** 14 female dancers (mean age 25.5 years, SD 3.7) participated in the study. 10 dancers recorded positive scores on the dance health questionnaire, suggesting a low risk of relative energy deficiency in sport (RED-S). 2 dancers were taking hormonal contraception. Apart from 1 dancer, those not on hormonal contraception reported current eumenorrhoeic status. The initiative of monitoring menstrual cycles and application of AI to model menstrual cycle hormones found that subclinical hormone disruption was occurring in 6 of the 10 dancers reporting regular cycles. 4 of the 6 dancers who received personalised advice, showed improved menstrual hormone function, including one dancer who had planned pregnancy.

**Conclusions:** Multimodal monitoring facilitated delivery of prompt personalised clinical medical feedback specific for dance. This strategy enabled the early identification and swift management of emergent clinical issues. These innovations received positive feedback from the dancers.

**Summary boxes:** *What are the new findings?:*

- Monitoring female dancers with a variety of interactive methods – dance specific questionnaire, online tracking and blood testing – together with individual clinical discussion, facilitates comprehensive, personalised support for dancer health.
- The clinical application of artificial intelligence (AI) techniques to endocrine function provides the finer detail of female hormone network function.
- This novel approach to monitoring dynamic hormone function enabled the detection of subtle female hormone dysfunction as a result of changes in training and nutrition patterns, which occurred before change in menstruation pattern from menstrual tracking.
- This multifaceted clinical approach was also effective and helpful in supporting dancers restore full hormone network function through personalised training and nutritional strategies.

*How might this study impact on clinical practice in the future?:*

- Personalised, dance specific health advice based on subjective and objective measures can support sustainable individual dancer health.
- Clinical application of artificial intelligence (AI) to menstrual cycle hormones can provide a dynamic and complete picture of hormone network function, without the need to do daily blood tests to measure all four key menstrual cycle hormones.
- This AI approach to modelling hormones enables early detection of subtle, subclinical endocrine dysfunction due to low energy availability in female exercisers. This clinical tool can also facilitate the close clinical monitoring of the restoration of full hormone network function in recovery from low energy availability.
- Using AI to model female hormones can be an important clinical tool for female athletes, including those athletes where it is difficult to distinguish between perimenopause symptoms and those associated with low energy availability.

## Introduction

Professional dancers have a demanding schedule from a physical and psychological point of view and unlike athletes, there is no clear off-season of reduced training and performance. Furthermore, when combined with the aesthetic requirements of dance, dancers can be at risk of low energy availability, whether intentional or unintentional, and of developing the adverse clinical and performance outcomes of relative energy deficiency in sport (RED-S)[1].

In the clinical setting, a RED-S diagnosis is confirmed from clinical history, including menstrual status in females and blood testing to exclude medical causes. These clinical assessments, together with dual-energy X-ray absorptiometry scans to assess bone health can be used for risk stratification[2]. However, these metrics only become valuable once the adverse outcomes of low energy availability have occurred. Early detection of exercisers at risk of low energy availability is crucial in order to provide interventions to prevent progression to RED-S.

A recently published dance energy availability questionnaire (DEAQ) identified indicators and correlates of low energy availability and provided a RED-S risk score, allowing it to be used as a screening questionnaire for male and female dancers[3]. In terms of blood testing, for female athletes, even for those with functional hypothalamic amenorrhoea (FHA), all the key menstrual cycle hormones, follicle stimulating hormone FSH, luteinising hormone LH, oestradiol and progesterone, can be within the follicular range, albeit towards the lower end. Furthermore, it is challenging to identify subtle hormone network dysfunction outside of the research setting, despite high incidence of reported subclinical luteal phase deficits[4], which can have adverse health consequences. This is because there are constraints on the practicality and budget of serial blood testing, which is required to measure all four female hormones over a menstrual cycle. Similarly, for female athletes recovering from low energy availability, there is no practical way to monitor hormone network recovery to guide behaviours to restore energy availability, which can lead to a high dropout rate[5]. The challenge is to gain the maximum insights from the minimal amount of data. Artificial intelligence techniques are advocated and being shown to be valuable in personalisation of medicine[6].

The objectives of this study were to examine the value of health monitoring through a variety of modalities and analyses in professional dancers, including the clinical application of AI techniques to modelling menstrual cycle hormones.

## Methods

### Study design

A longitudinal study of professional female dancers at a dance company to monitor health through a variety of monitoring methods and analyses. All participants provided informed consent.

### Recruitment

Professional adult female dancers at a dance company were invited to participate from March 2020 through to July 2021

### Dance specific health questionnaire

An online dance specific health questionnaire was completed by all the dancers and a RED-S risk score was calculated, as previously published[14]. Virtual clinical interviews were conducted by Dr Keay with individual dancers to clarify any areas from the questionnaire.

### Online monitoring

Dancers logged daily wellbeing and training metrics. An additional menstrual tracking feature was developed.

### Blood biomarkers

Blood biomarkers were assessed for those female dancers not on hormonal contraception and menstruating, based on capillary blood samples, taken on day 14 and day 21 of a menstrual cycle. Samples were analysed for FSH (follicle stimulating hormone), LH (luteinising hormone), oestradiol and progesterone, at Surrey University accredited laboratories using a cobas8000 machine.

### Clinical interview

Initial individual dancer discussion was based on the responses to the dance heath questionnaire. Regular virtual follow-up occurred to discuss blood test results, logged wellbeing and menstrual cycle regularity and/or issues. Personalised dance specific actionable advice was provided, with regard to dancer behaviours, including nutrition and training load.

### Dancer feedback

Feedback was sought on the initiatives of multimodal health monitoring and on demand virtual clinical input.

### Statistical analysis

Data analysis was performed using the open-source tools, SciPy and Pandas (NumFOCUS, Austin, Texas), implemented using the Python programming language. Summary statistics, including count, mean and standard deviation of responses, were calculated for the overall sample and by subgroup, according to the responses to the questions. Body mass index (BMI) was calculated by dividing weight (kg) by the square of height (m). Minimum BMI (BMI min) was calculated based on the dancer’s minimum weight for current height. A weight variability variable was calculated for the current height by dividing the difference between the maximum and minimum weights by the current weight.

The modelling of the time variation of female hormones drew from the field of Artificial Intelligence. A tested system used Bayesian Inference to infer the best fit curves of the four key menstrual cycle hormones. The analysis was performed for each individual, taking into account reported cycle length and the measured hormones at two time points in the menstrual cycle. The output was used to assess overall network function.

### Participant involvement

Participant involvement was integral at all stages of this research. Dancer feedback on monitoring systems and rapid virtual clinical access was sought and recorded with a follow up feedback questionnaire.

## Results

14 professional female dancers participated in the study (mean age 25.5 years, SD 3.7)

### Dancer Health Questionnaire

Figure 1 shows that the questionnaire results were consistent with a reasonably healthy disposition across the group, with no dancers producing strongly negative RED-S Risk Scores, in comparison with the broader study. RED-S Risk Scores from the previous dancer study fell in the range of -15 to +15, with negative scores being indicative of low energy availability. None of the professional dancers were particularly extreme relative to the broader study of younger dancers. Four of the dancers showed notably negative scores, each exhibiting two of more of the typical combination of low minimum BMI (<19), disrupted menstrual function and strong desire to control weight and food.

**Figure 1:**
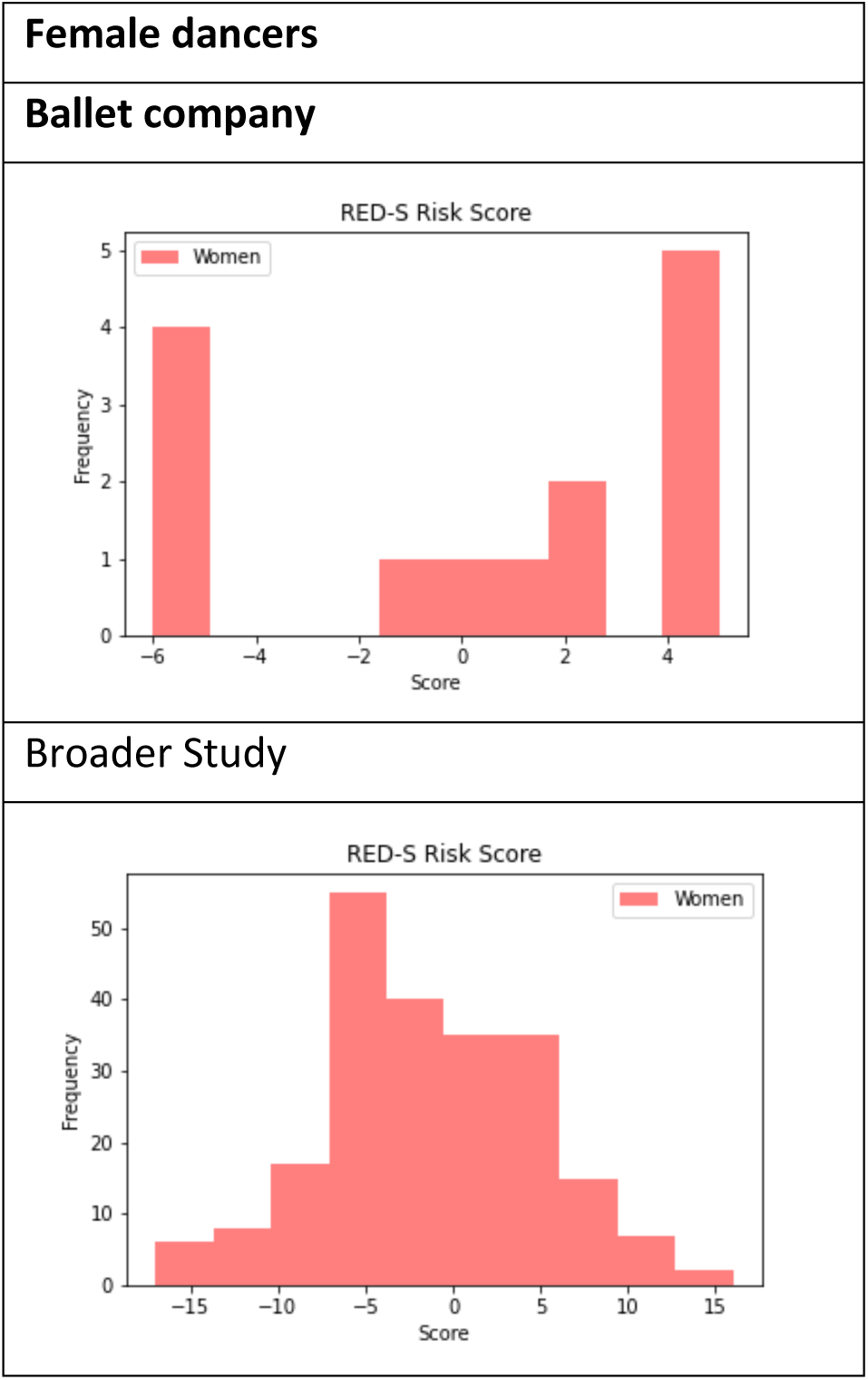
RED-S Risk Scores for company dancers versus previous dance study

### Hours of training

Table 1 shows that the company dancers complete a large number of hours of exercise per week, with the total ranging from 26 to 60 hours. Even during restrictions to dance studios early in the COVID-19 pandemic, dancers maintained high levels of training with virtual classes and other training modalities.

**Table 1:**
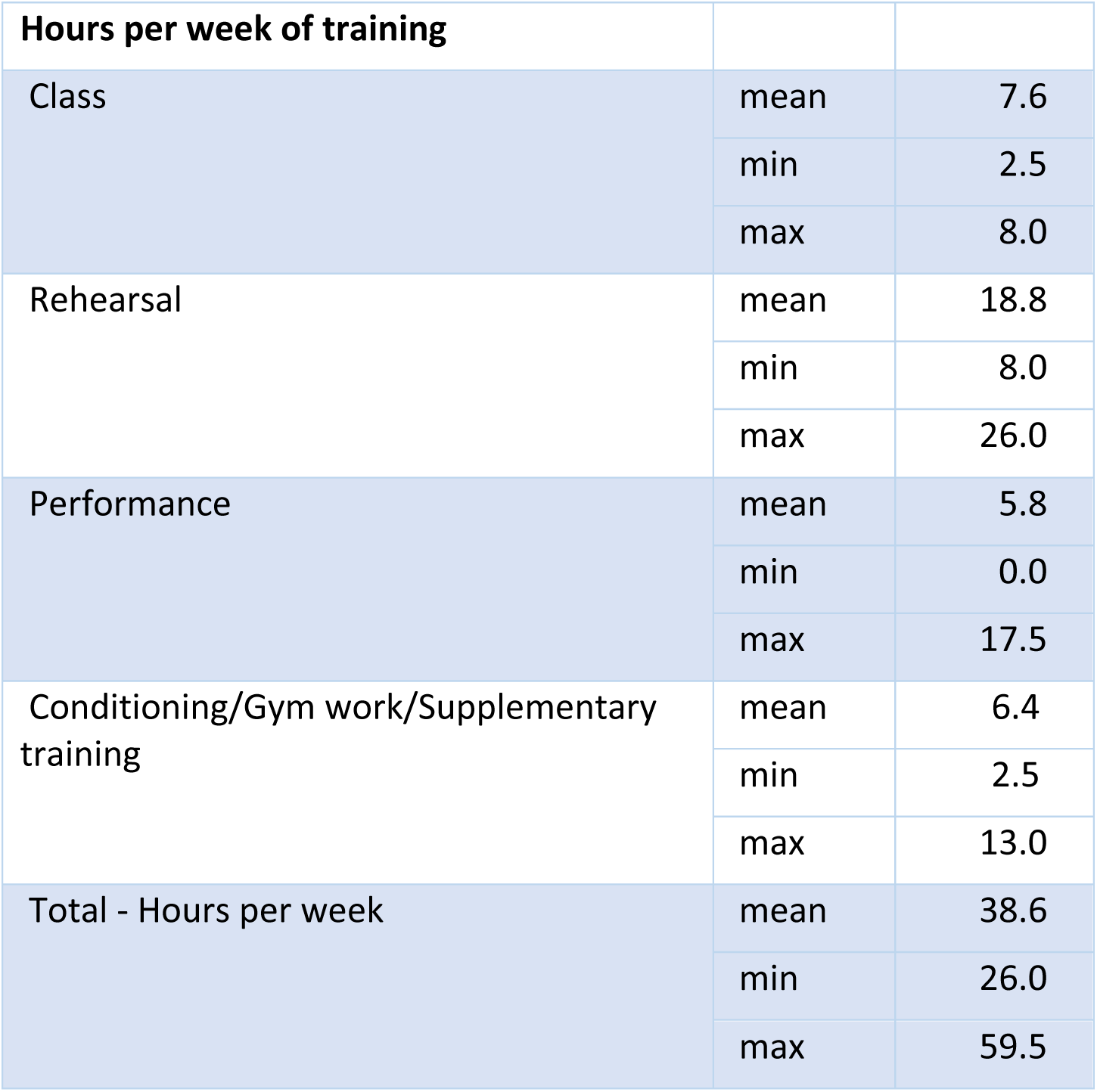
Weekly hours of training

### Anthropomorphic data

**Table 2:** Anthropometric data for the dancers

| Anthropometric data |  |  |
| --- | --- | --- |
| BMI | mean | 19.5 |
|  | std | 1.0 |
|  | min | 18.1 |
|  | max | 21.5 |
| Current height (in metres) | mean | 1.70 |
|  | std | 0.1 |
|  | min | 1.6 |
|  | max | 1.8 |
| Current weight (kg) | mean | 55.3 |
|  | std | 5.0 |
|  | min | 47.0 |
|  | max | 62.0 |

### Wellbeing and psychological report

Dancers were asked to score on scale of 1 to 5 wellbeing metrics relating to freshness, sleep quality and digestive system function and results are shown in Table 3. The mean score for dancers was mid-range.

**Table 3:** Dancer wellbeing scores

| Wellbeing scores |  |  |
| --- | --- | --- |
| How would you rate your levels of freshness over the past year? | mean | 2.8 |
|  | min | 2.0 |
|  | max | 4.0 |
| How would you rate your sleep quality over the past year? | mean | 3.1 |
|  | min | 1.0 |
|  | max | 5.0 |
| How would you rate your digestive system over the past year? | mean | 3.2 |
|  | min | 1.0 |
|  | max | 5.0 |

Regarding psychological factors, in response to the question “How do you feel if you have to miss class/rehearsal?”: 9 responded that “these things happen” and 5 reported feeling anxious.

Table 4 shows the scores for rating importance of controlling food intake and weight. The mean score indicates a relaxed attitude

**Table 4:** Scores for rating importance of control aspects

| Control scores |  |  |
| --- | --- | --- |
| Controlling what you eat | mean | 2.7 |
|  | min | 0.0 |
|  | max | 4.0 |
| Controlling what you weigh | mean | 2.1 |
|  | min | 0.0 |
|  | max | 5.0 |

### Menstrual function

Seven of the fourteen female dancers had experienced amenorrhoea in the past as students in training. However, currently the ten dancers not on hormonal contraception reported menstruation with more than 9 periods per calendar year.

### Injury and illness

The dancers had experienced a relatively low level of injury and illness, with just one or two experiencing longer periods off.

## Monitoring

### Wellness, training load, injury

During the spring 2020 lock down due to the COVID-19 pandemic at the start of the study, no access to dance studios for training or theatres for performance was possible. Nevertheless, some dancers reported an overall increase in training load with access to numerous online dance classes and supplementary exercise. The return to the studios in late summer was on a reduced schedule, in order to keep dancers in support bubbles, nevertheless with the rapid increase in rehearsal time, the injury rate slightly increased in autumn.

### Menstrual monitoring

A menstrual tracking facility was successfully integrated into the existing monitoring system for dancers, enabling female dancers to track all training metrics in one place, which was well received. From the clinical perspective, this was also made remote monitoring possible, which proved particularly useful during lockdown. Modifications were made based on invited feedback from dancers to provide information about menstrual cycles.

During the initial spiring lockdown a few dancers reported slight disruption of cycles. Despite the reduced training load, as there was no access to the studios, many dancers reported that change in routine was challenging. Some dancers expecting to perform in US had to fly home, with some dancers reporting symptoms consistent with COVID-19.

With a return to the studio, albeit with a reduced training schedule, dancers, who had experienced some degree of menstrual disruption, reported resumed regularity of periods, although one dancer reported lengthened cycles associated with continuation of out of studio exercise.

Follow up feedback on this aspect of the study was rated highly by the dancers who cited that this was a helpful addition to current logging of wellbeing and training load.

### Female hormone network modelling

This was performed for the ten female dancers not taking hormonal contraception, with a total of 17 menstrual cycle tests. Eleven of these tests showed subclinical downregulation of the hypothalamic network, indicated by low range variation of FSH, LH, oestradiol and progesterone, in spite of menstrual tracking displaying normal regularity. This coincided with the dancers reporting the challenges of changed and uncertain routine with initial lock down. Although not able to go into the studio, these particular dancers reported an increase in exercise to compensate, coupled with concern about appropriate nutritional intake. Six dancers completed more than one menstrual cycle test, with three dancers showing improved hormone function after receiving personalised clinical advice about nutrition and training load. One of the dancers, who performed a single cycle test indicating probable ovulation, went on to become pregnant.

Follow up feedback on this aspect of the study was rated with the highest score by all dancers who cited that the personalised hormone information and explanation was very interesting and helpful.

### Clinical Interviews

Virtual individual dancer discussion revealed that dancers were finding the situation challenging of not being able to access the company building for either training or on person clinical support. Having the opportunity to discuss results and explore strategies to remain healthy and fit was particularly welcomed during the pandemic. Similarly, returning to the studio, together with the opportunity to discuss strategies was reported as being helpful. All the dancers rated this aspect of the study with the highest score, indicating that personal discussion with a medical doctor familiar with dance demands was extremely valuable.

## Discussion

The results from dancer health questionnaire and serial, static blood testing showed that the majority of dancers were in good health. For female dancers the addition of menstrual cycle tracking to the company monitoring system was effective at highlighting any issues. The application of AI techniques to modelling menstrual cycle hormones provided important clinical detail to personalise clinical advice to avert and support recovery from low energy availability.

### Dance Specific Health Questionnaire

As reported previously, activity specific questionnaires are effective in identifying those displaying psychological, physical and physiological indicators of low energy availability[3]. This is a practical clinical tool to help offer swift support to those at risk of developing adverse clinical outcomes of RED-S. The health questionnaire scores from the professional dance company were better than the previous study focused on pre-professional dancers. Most likely this reflects the fact that dancers have to be in good health in order to secure and maintain professional contracts. This is supported by the finding that although a few female company dancers had experienced brief episodes of secondary amenorrhoea as vocational students in full time training, all the dancers in the current study, not on hormonal contraception, were menstruating. Encouragingly, most dancers reported taking vitamin D supplementation and had good levels of vitamin D, which has been shown to support dancer health and performance[7].

Using this dance specific questionnaire as part of the onboarding process for dancers joining a dance company would be helpful and also as a yearly quantitative monitoring tool to support dancer health. Nevertheless, a recent review of questionnaires for low energy availability found that these were not so effective in assessing unintentional low energy availability with increased energy expenditure[8].

### Dancer subjective monitoring

The dancers in the company had a monitoring system to log daily well-being and training load. Nevertheless, a more personalised training metric might be provided through blood biomarker tracking as hormones are required to drive adaptive responses to exercise. In general, there was a good match between subjective reports and blood testing results.

### Monitoring female hormone network function: menstrual tracking

Female hormone networks are particularly sensitive to low energy availability[9]. The mechanism of hormone disruption caused by low energy availability is down regulation of the hypothalamic-pituitary axis. Cumulative low energy availability can result in functional hypothalamic amenorrhoea (FHA). As there are health and performance consequences of FHA, it is a clinical priority to identify those at risk[10]. For those not taking hormonal contraception, menstrual cycle tracking is a very good starting point and was successfully integrated into the dancer health monitoring in the company alongside daily well-being metrics and training load. Dancer feedback was important to make this menstrual cycle tracking aspect easy to use. The dancers reported that having a menstrual tracking system with tips for dealing with any issues, alongside monitoring of wellbeing and training load was helpful. As a result the dancers at the conclusion of this study did not report that menstrual cycles disrupted dance training, which contrasts to a recent study of fifteen female rugby players where 67% reported menstrual cycle issues impacted training[11]. Having this tracking facility alongside pre-existing monitoring functionality was also practical and helpful for healthcare staff, especially when face-to-face meetings were not possible during the pandemic. This new monitoring was highly rated by the dancers from the feedback survey.

### Monitoring female hormone network function: artificial intelligence

In the absence of measuring hormone levels, menstrual tracking alone can miss subclinical anovulatory cycles, where periods are regular, yet suboptimal levels of ovarian hormones can have adverse health effects[12]. This was the case in some dancers in this study, especially during the initial lockdown. Unlike urine or saliva sampling, a blood test measures of all the female hormones (FSH, LH, oestradiol and progesterone). Nevertheless, a single blood test, which is typically performed in the early follicular phase, only provides information at a single point in time. While daily blood sampling would be expensive and impractical, the use of AI techniques to model hormonal variation for menstruating females, gives rise to the convenient alternative of taking two capillary blood samples over a menstrual cycle.

Early detection of the impact of low energy availability on female hormone networks is important from a clinical point of view. Low fluctuating levels of ovarian hormones can have adverse health effects per se and in sustained low energy availability this situation can lead to complete down-regulation of the hypothalamic-pituitary-ovarian axis found in FHA. The adverse health and performance consequences of FHA are well described in the clinical syndrome of RED-S. Furthermore, specifically in female dancers, there is evidence of long-term irrecoverable effects of FHA on bone health from a study of retired dancers[13]

Therefore, early identification of athletes in low energy availability is important in the prevention of progression to RED-S. The effectiveness of an exercise specific questionnaire, combined with clinical interview has been shown to be effective in detecting male athletes at risk of RED-S[14]. Whilst there is a validated screening questionnaires for female athletes, this is not exercise specific[15]. Furthermore, as shown in this study and other studies of exercising females, report of regular menstrual cycles may belie subtle menstrual hormone dysfunction[16]. Low energy availability is caused by a mismatch of energy intake and energy expenditure which can be intentional or unintentional[17]. Objective endocrine assessment of this situation is of clinical value. If identification of low energy availability is timely, prompt clinical management to support behavioural change, has been demonstrated to be effective in restoring health and athletic performance of male athletes[18].

In female athletes the recovery from menstrual disruption due to low energy availability, the magnitude of energy intake is not known[5]. Most likely this is due to individual differences between females. Resumption of regular periods is an encouraging clinical sign, nevertheless it is not possible to be certain that full hormone network function has been restored. Having quantified detail about the effect of non-pharmacological strategies on recovery of menstrual cycle hormone function would be very valuable[19].This study found that modelling all the menstrual cycle hormones over an entire cycle for an individual female proved a personalised, practical and sensitive clinical tool to indirectly monitoring energy availability. Dancers identified as having subclinical endocrine network dysfunction, were promptly provided with personalised advice on nutrition timing and training load. Subsequent checking full endocrine network restoration was helpful for these dancer and clinician to give feedback that personalised behavioural change had been effective and so support effective change in the long term. Equally for those dancers who did not restore full endocrine function, then further personalised advice around training and nutrition could be targeted and endocrine response quantified.

From a clinical medical point of view, having an integrated system using AI proved an effective way to check in on female dancers and address any issues promptly. This provided real-time monitoring to facilitate proactive rather than reactive action.

From the point of view of the professional female dancers, all participants gave this integrated, personalised hormone health information and explanation the highest feedback score. Dancers cited the value of this personalised hormone innovation for their health and performance.

### Limitations and further work

Although the dancers all came from the same dance company, self-selection bias could play a part in terms of those interested in menstrual tracking and hormone modelling. Further work will be to include older dancers such as teaching and artistic staff where it is currently difficult to distinguish between the effects of low energy availability and physiological changes in endocrine networks, such as declining ovarian response occurring during perimenopause in lead up to menopause. Other avenues are to develop similar AI approaches specific for male athletes as a practical quantitative tool in the early identification of low energy availability and in monitoring of effective restoration of endocrine networks with non-pharmacological, behavioural interventions.

## Conclusions

The combination of a dance specific questionnaire screening tool, longitudinal monitoring of clinical biometric measures with application of AI techniques on female hormone networks and prompt personal clinical discussion were initiatives successful in identifying and personalising clinical advice. This approach was well received by professional dancers.

## Data Availability

Data available on reasonabale request

## Statements

## Acknowledgments

Thank you to the company artistic and healthcare staff and all the dancers for their interest and participation in this study.

## Contributors

NK, ML, GF: conceptualisation of project, development of study design, collation of data, data analysis (GF), data interpretation (GF, NK), drafting and revision of manuscript.

## Data sharing

No unpublished data were used in the preparation of this manuscript

## Ethical Approval

This study was reviewed and approved by Durham University research ethics committee.

